# Comparative Evaluation of Cross-Ancestry Polygenic Risk Scoring of Type 1 Diabetes in the All of Us Cohort

**DOI:** 10.1101/2025.10.02.25337098

**Authors:** Mohammad Ahangari, Spencer Moore, Ivan Davidson, Jonathan Anomaly, Robert Maier, Jeremiah H. Li, Michael Christensen, David Stern, Tobias Wolfram

**Affiliations:** Herasight Research, USA

**Author notes:** Contributed equally (David Stern, Tobias Wolfram).

## Abstract

Type 1 diabetes is a highly heritable autoimmune condition characterized by the destruction of pancreatic beta cells, resulting in insulin deficiency. Here, we developed a novel polygenic score we call the HLA-Augmented SBayesRC Framework (HLA-ARC). HLA-ARC integrates direct modeling of HLA haplotypes, with a Bayesian regression approach for the non-HLA component. SBayesRC leverages extensive functional genomic annotations and linkage disequilibrium patterns across approximately 7.4 million variants, substantially enhancing predictive accuracy. We systematically compared HLA-ARC to three existing T1D polygenic scores (Polygenic Risk Score extension for Diabetes Mellitus [PRSedm], Trans-Ancestry Polygenic Score for Diabetes [TA-PS], and Type 1 Diabetes Multi-Ancestry Polygenic Score [T1D-MAPS]) using data from the ancestrally-diverse All of Us cohort. Among the three existing methods, T1D-MAPS showed superior performance in all ancestry groups. However, HLA-ARC consistently outperformed the existing methods, achieving AUROC values exceeding 0.91 in European individuals and 0.89 in non-European groups. Our results demonstrate that integrating HLA haplotype modeling with genomic annotation and ancestry-informed linkage disequilibrium methods significantly improves polygenic risk prediction for autoimmune diseases characterized by major genetic risk loci.

## Introduction

Type 1 diabetes (T1D) is a highly heritable autoimmune disease characterized by the destruction of pancreatic beta cells and absolute insulin deficiency, with substantial genetic risk conferred by variants in the human leukocyte antigen (HLA) region on chromosome 6. The HLA region encodes proteins essential for immune recognition and antigen presentation. Although the life- time risk of T1D is approximately 0.4-0.6% in the general population, it increases significantly to around 6% for individuals with an affected dizygotic twin, and to 25–50% for those with an affected monozygotic twin ^1^.

The HLA region accounts for a large fraction of T1D heritability ^2^, with higher risk conferred by specific HLA class II haplotypes. The DR4-DQ8 and DR3-DQ2 haplotype combination demonstrates strong associations, with odds ratios reaching *∼*15 for T1D susceptibility ^3, 4^. These haplotypes and their specific combinations, influence T1D risk by determining the peptide-binding specificity of HLA class II molecules, which enhances presentation of beta cell autoantigens and facilitates the breakdown of immune tolerance.

Beyond the HLA region, genome-wide association studies (GWAS) have identified over 50 significant non-HLA loci that modify disease risk, each contributing relatively modest effects to T1D susceptibility ^5^. This genetic architecture, dominated by a few high-effect HLA haplotypes with additional contribution from many small-effect variants, poses distinct challenges for constructing accurate polygenic scores (PGS).

Conventional genome-wide scoring methods typically treat all loci uniformly, failing to account for the outsized contribution of the HLA region in T1D. This limitation is compounded across diverse populations where HLA haplotype frequencies and LD patterns differ substantially. For instance, European-derived T1D polygenic scores have substantially reduced predictive accuracy when applied to non-European populations, particularly when HLA-specific risks are not explicitly modeled ^6^. This discrepancy underscores the need for ancestry-aware modeling frameworks that properly handle both HLA and genome-wide variation across ancestries.

In this study, we introduce the HLA-Augmented SBayesRC Framework (HLA-ARC), an extension of T1D-MAPS ^7^ designed to further refine the non-HLA component of T1D PGS by integrating LD information and functional annotations for approximately 7.4 million genetic markers, using the Bayesian PGS construction method SBayesRC ^8^. SBayesRC has demonstrated enhanced cross-ancestry predictive accuracy, achieving improvements of up to *∼* 34% compared to its predecessor, SBayesR ^9^. HLA-ARC combines direct genotyping and phased scoring of high-risk HLA DRB1-DQA1-DQB1 haplotypes with the genome-wide non-HLA component derived from SBayesRC, resulting in a unified, ancestry-informed PGS (Figure 1). Notably, HLA-ARC can be extended to other diseases that require careful modeling of both HLA and non-HLA components, such as multiple sclerosis and other autoimmune conditions.

**Figure 1.**
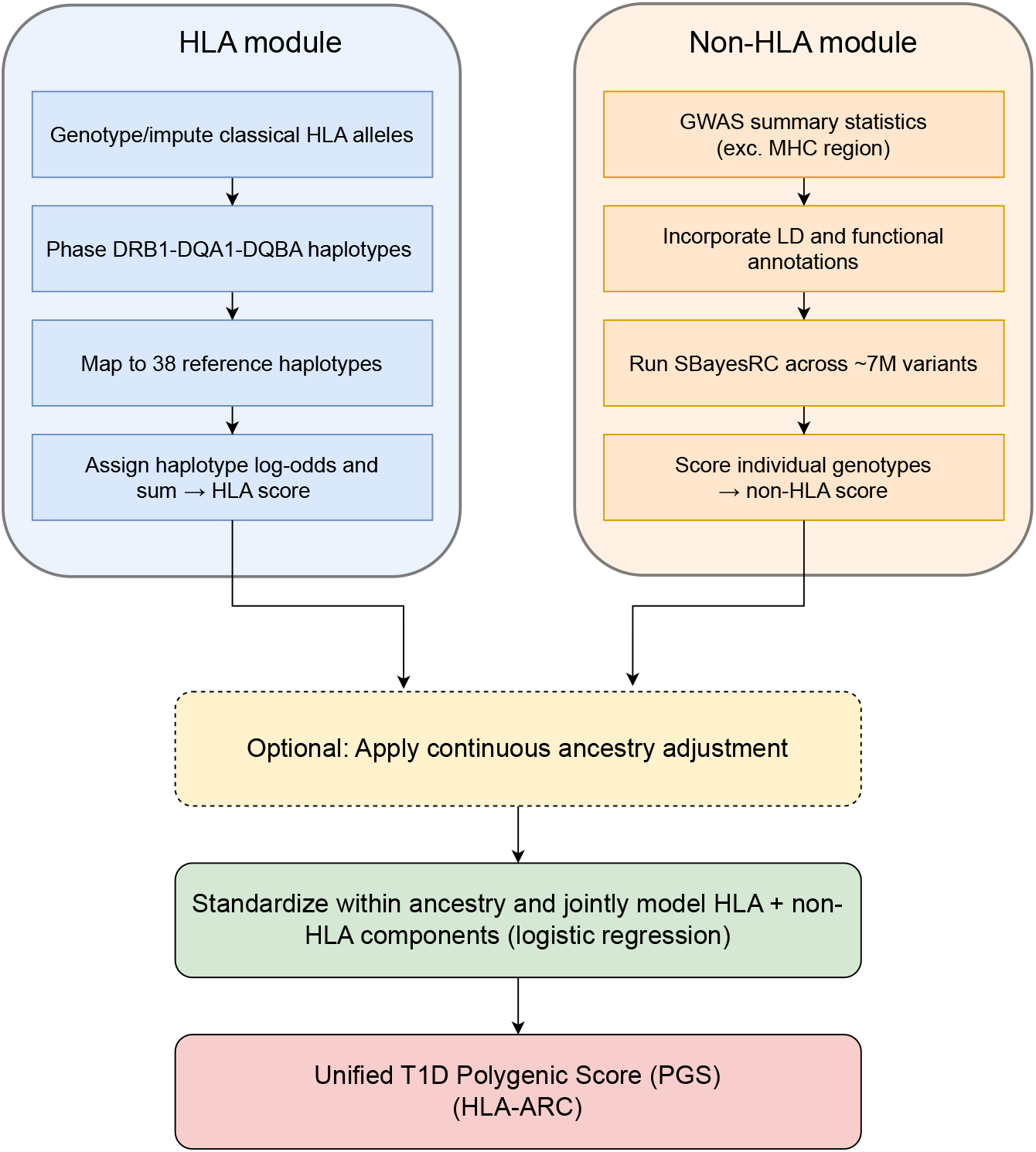
HLA-ARC workflow. HLA alleles are directly genotyped or imputed and scored as phased DRB1 DQA1 DQB1 haplotypes. The non-HLA variants are scored using SBayesRC posterior effects estimated across ∼7.4M variants with LD and functional annotations. The two standardized components are combined with a joint model to produce unified T1D PGS weights. The two components are then jointly modeled to generate the unified PGS.

We find HLA-ARC attained meaningful risk stratification between T1D cases and controls across diverse ancestry groups using data from the All of Us (AoU) cohort ^10^, comprising over 400,000 individuals with whole-genome sequencing data. We then systematically benchmarked HLA-ARC against three published state-of-the-art PGS frameworks – PRSedm ^11^, TA-PS ^12^, and T1D-MAPS – and found that HLA-ARC outperformed all tested methods across all ancestries. HLA-ARC thus provides a more accurate and equitable framework for T1D genetic risk assessment across ancestry groups, surpassing current state-of-the-art methods in predictive performance.

## Results

### Study populations and modeling overview

All models were evaluated in the All of Us (AoU) Research Program using strict EHR-based T1D definitions. After quality control and phenotype curation, AoU provided 200 T1D cases and 21,802 controls (EUR *n* = 78/7,716; AFR *n* = 54/4,029; AMR *n* = 51/6,462; other ancestries were under-powered for ancestry-specific primary analyses). Classical HLA alleles (DRB1, DQA1, DQB1) were inferred and phased to haplotypes; the non-HLA component of the score was constructed using SBayesRC applied to *∼*7.4M variants. Both components were then combined in a weighted sum to compute individual scores (see Methods).

### HLA-ARC score distributions highlight T1D risk stratification and provide practical risk thresholds

Kernel-density plots of the HLA-ARC scores with continuous ancestry adjustment show clear case-control stratification within each ancestry group (Figure 2A). The right-shifted distributions in T1D cases relative to controls are most pronounced in EUR and remain distinct in AFR and AMR, as well as combined cohorts. Using the equal-density threshold (shown as a dashed line), we demonstrate robust discrimination, but also highlight slightly reduced separation in nonEUR groups, with sensitivity/specificity of 0.84/0.89 in EUR and 0.77/0.86 in non-EUR cohorts, respectively. Figure 2B displays AUROC curves from continuous ancestry-adjusted HLA-ARC scores (see Methods). The left-skewed, rapidly rising profiles show that sensitivity is achievable at low false-positive rates across all ancestries studied, further supporting the portability of these scores across ancestries.

**Figure 2.**
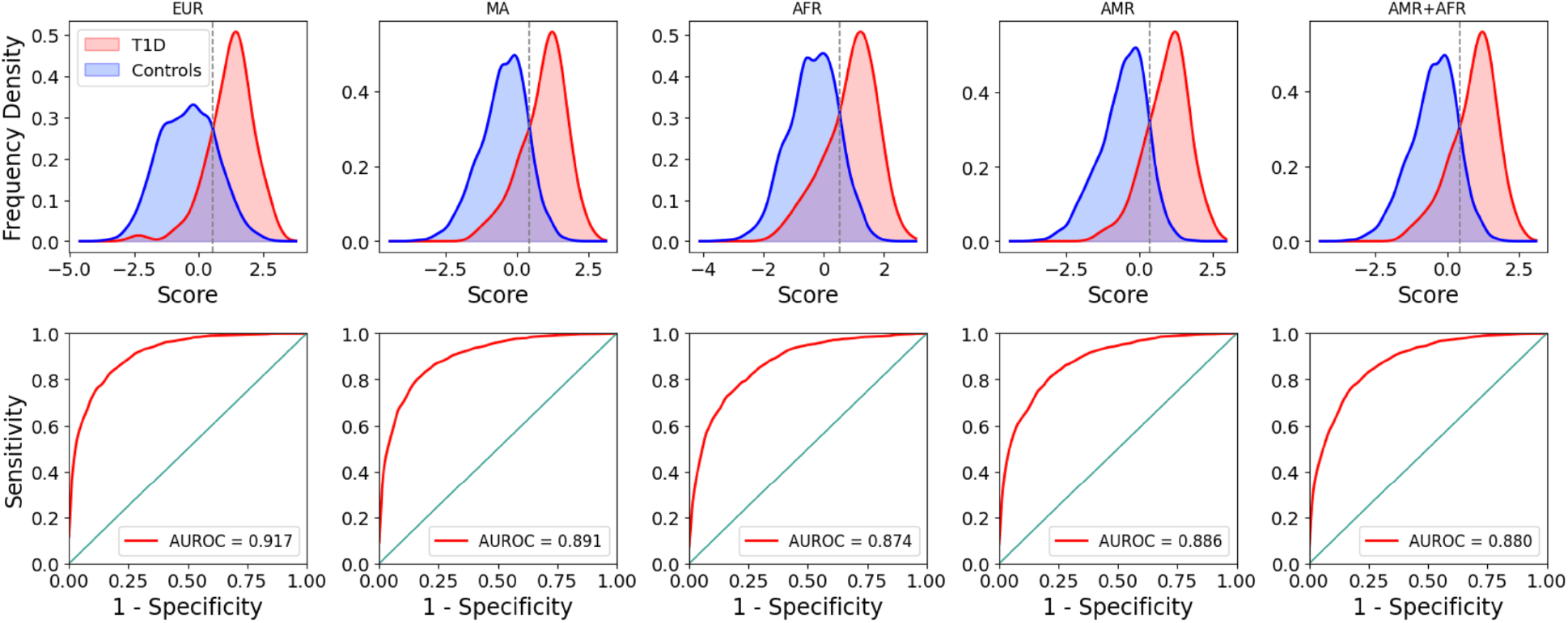
Ancestry-stratified performance of the HLA-ARC scores with continuous ancestry adjustment. (A) kernel-density estimates of cases (red) and controls (blue). The dashed line marks the equal-density threshold (intersection of KDEs). Reported operating points at this threshold were: EUR 0.84/0.89 (sensitivity/specificity), AFR 0.75/0.85, AMR 0.81/0.87, and AMR+AFR 0.77/0.86. (B) AUROC curves of the same scores, with diagonal lines indicating no discrimination null model and red lines showing the observed performance. Curves remain well above the diagonal across all ancestries, indicating robust discrimination.

### Comparison with three published approaches

In analyses run alongside established methods (Table 1), HLA-ARC matched or exceeded their performance (Figure 3). The standard PRSedm model showed moderate discrimination for T1D across ancestries in the AoU cohort, consistent with prior literature ^7^. In the EUR subset, PRSedm achieved an AUROC of *∼*0.882, whereas performance was notably lower in the AFR subset (*∼*0.784) and intermediate in the AMR group (*∼*0.793). Correspondingly, variance explained on the liability scale by PRSedm was modest across all groups, with the lowest (0.189) observed in AFR individuals.

**Table 1:** Summary of the PGS methods compared in this study, highlighting how each approach handles HLA and non-HLA components of the PGS.

| Method | HLA component | non-HLA component | Score integration approach |
| --- | --- | --- | --- |
| **PRSedm** | Fixed set of 67 established variants | Limited to 32 non-HLA markers | Unweighted linear sum |
| **TA-PS** | Fixed set of 67 established variants | 1.2M non-HLA with PRS-CS | Unweighted linear sum |
| **T1D-MAPS** | Direct genotyping of DRB1, DQA1, and DQB1 | 1.2M non-HLA markers with PRS-CS | Weighted linear combination |
| **HLA-ARC** | Direct genotyping of DRB1, DQA1, and DQB1 | 7.4M non-HLA markers with SBayesRC | Weighted linear combination |

**Figure 3.**
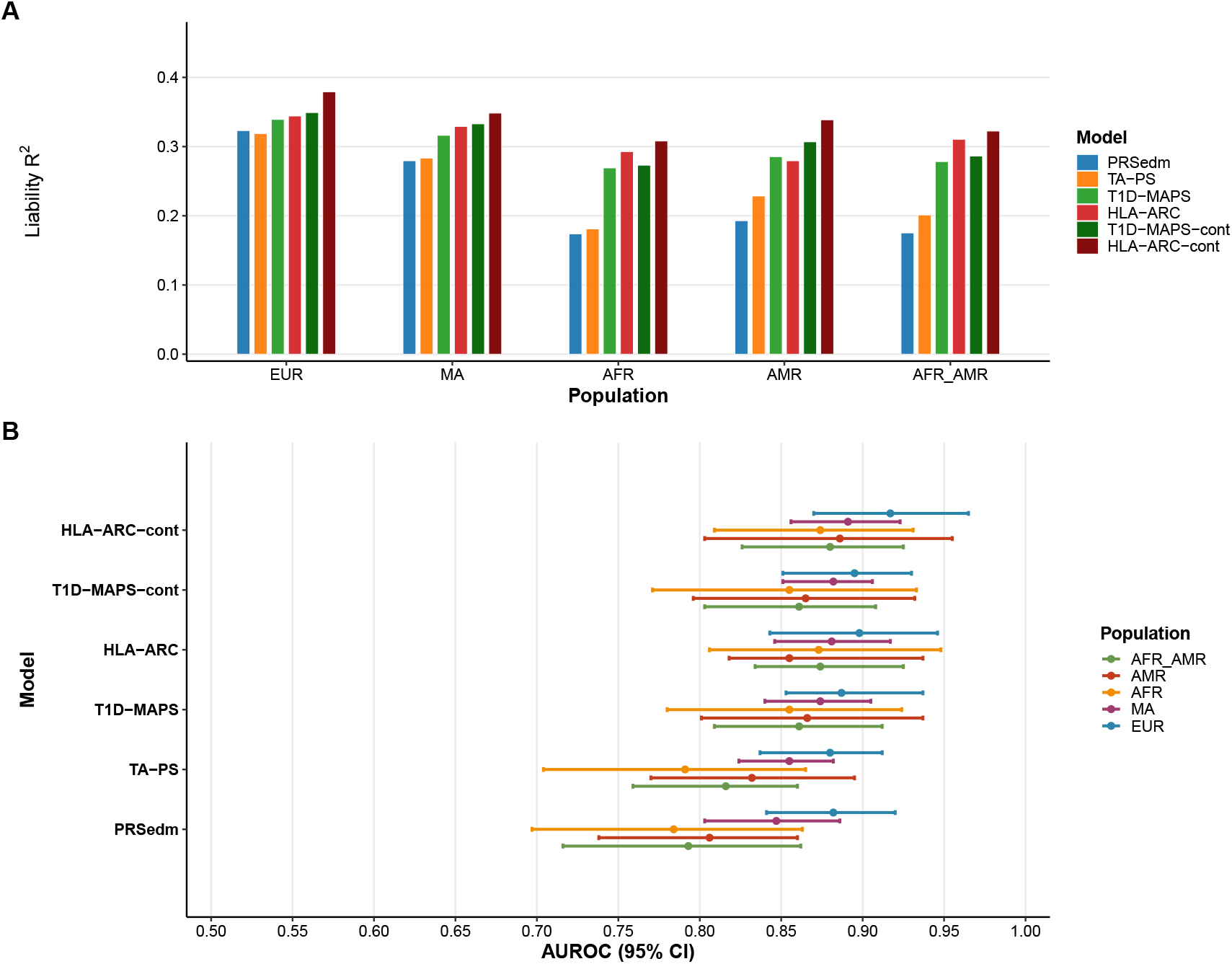
Performance of polygenic risk score models for Type 1 Diabetes across diverse ancestries. (A)Liability-scale R^2^ values for six T1D prediction models across five ancestry groups (EUR: European, MA: Middle Eastern, AFR: African, AMR: American, AFR_AMR: African American). (B) Area under the receiver operating characteristic curve (AUROC) with 95% confidence intervals for the same models.

Incorporating genome-wide PGS alongside the HLA component improved predictions relative to PRSedm. TA-PS, which uses PRS-CS-derived weights ^13^, raised AUROC in AFR and AMR ancestries (e.g., from *∼*0.80 to above 0.83 in AMR), with EUR performance remaining around *∼*0.88. Overall, including genome-wide TA-PS data increased the fraction of T1D liability variance explained compared to PRSedm.

T1D-MAPS had superior predictive accuracy compared to PRSedm and TA-PS across all ancestries (Figure 3), with AUROC increasing from 0.78 to 0.85 in T1D in AFR ancestry. Adopting the HLA-ARC approach with comprehensive SBayesRC weights further enhanced discrimination, achieving AUROC values around 0.90 in EUR and more than 0.87 in non-EUR individuals (Figure 3). Consistent with this high prediction accuracy and discrimination power, HLA-ARC explained the largest proportion of liability variance across ancestries.

The relative predictive power of the HLA and non-HLA component may differ with ancestry. Specifically, we have previously shown that our SBayesRC scoring approach has reduced performance in ancestries that scales with the genetic distance of the test set of individuals from the training set ^14^. Thus, we applied a continuous ancestry adjustment to the SBayesRC component of the score, as the dispersed polygenic signal likely more closely obeys the ancestry attenuation patterns observed across polygenic traits, whereas the HLA alleles would not necessarily follow the same effect size attenuation pattern. As expected, we observed ancestry-dependent reduction in the effect sizes of both the HLA and non-HLA components when jointly regressed of T1D status that scaled proportionally to ancestral distance. Notably, the *relative* strength of the associations of the HLA and non-HLA components increased with genetic ancestry. The ratio of the log odds ratio of the HLA to non-HLA components increased from 1.46 in EUR to 2.02 in AMR and 2.57 in AFR, where notably the non-HLA component was not significantly associated with T1D status (Table 2), underscoring the importance of accurate HLA haplotyping in non-EUR individuals, and how HLA-driven risk is broadly comparable across populations once standardized on a common scale, whereas non-HLA effects attenuate in non-EUR groups. Applying continuous ancestry adjustments to account for the change in relative importance of the HLA and non-HLA components provided marginal gains for both T1D-MAPS and HLA-ARC. For example, HLAARC EUR scores slightly improved to *∼* 0.91, and non-EUR (AFR+AMR) scores increased from 0.87 to 0.88. These results suggest accounting for gradations of ancestry can modestly enhance prediction accuracy, particularly in non-EUR cohorts.

**Table 2:**
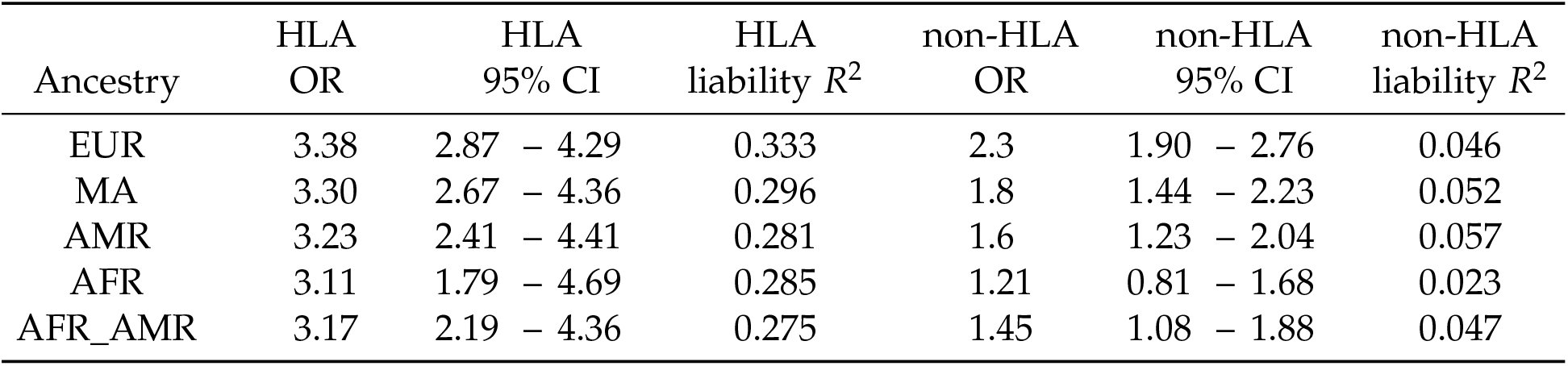
HLA-ARC-cont odds ratios (OR) with 95% confidence intervals (CI) and liability-scale *R*^2^ values for HLA and non-HLA components across ancestries. HLA consistently shows stronger effects and higher liability *R*^2^, while non-HLA effects attenuate in non-EUR groups.

### Using HLA-ARC in Preimplantation Genetic Testing

The high predictive accuracy of HLA-ARC across ancestries suggests potential utility in preimplantation genetic testing for polygenic traits (PGT-P). To evaluate this application, we developed a simulation framework modeling T1D risk reduction achievable through embryo screening (see Supplementary Methods).

Our approach models T1D liability as the sum of HLA effects, polygenic score, unmeasured genetic factors, and environmental variation. HLA haplotypes transmit via Mendelian inheritance, while the polygenic component exhibits within-family segregation variance of 1/2 per generation. Critically, we account for unmeasured familial factors that create within-family correlation—when parents are affected with T1D, their unmeasured genetic risk distribution is updated via Bayes’ rule based on their disease status. This allows us to model realistic scenarios where couples seek counseling without prior genotyping, with parental genetics treated as latent variables informed only by disease history.

We simulated 10,000 families across four parental history strata (Neither affected, mother/father affected, Both affected) for each ancestry (EUR, AMR, AFR) for a set of realistic, lower-bound estimates of the number of embryos available to screen (2-10) in line with previous literature ^15^. Three selection strategies were evaluated: random selection (baseline), selection based on measured genetics (HLA-ARC score, representing practical implementation), and oracle selection with access to all genetic and environmental factors (i.e. theoretical upper bound).

Figure 4 shows the remaining relative risk after HLA-ARC-based selection across ancestries and family histories. HLA-ARC-guided selection provides substantial risk reduction even when just two embryos are available for screening, achieving relative risk reductions (RRR) of approximately 42-51% in EUR, 32-38% in AFR, and 35-43% in AMR, depending on family history. RRR increases markedly with number of embryos: With 5 embryos screened RRR of 71-80% in EUR, 57-65% in AFR, and 61-71% in AMR.

**Figure 4.**
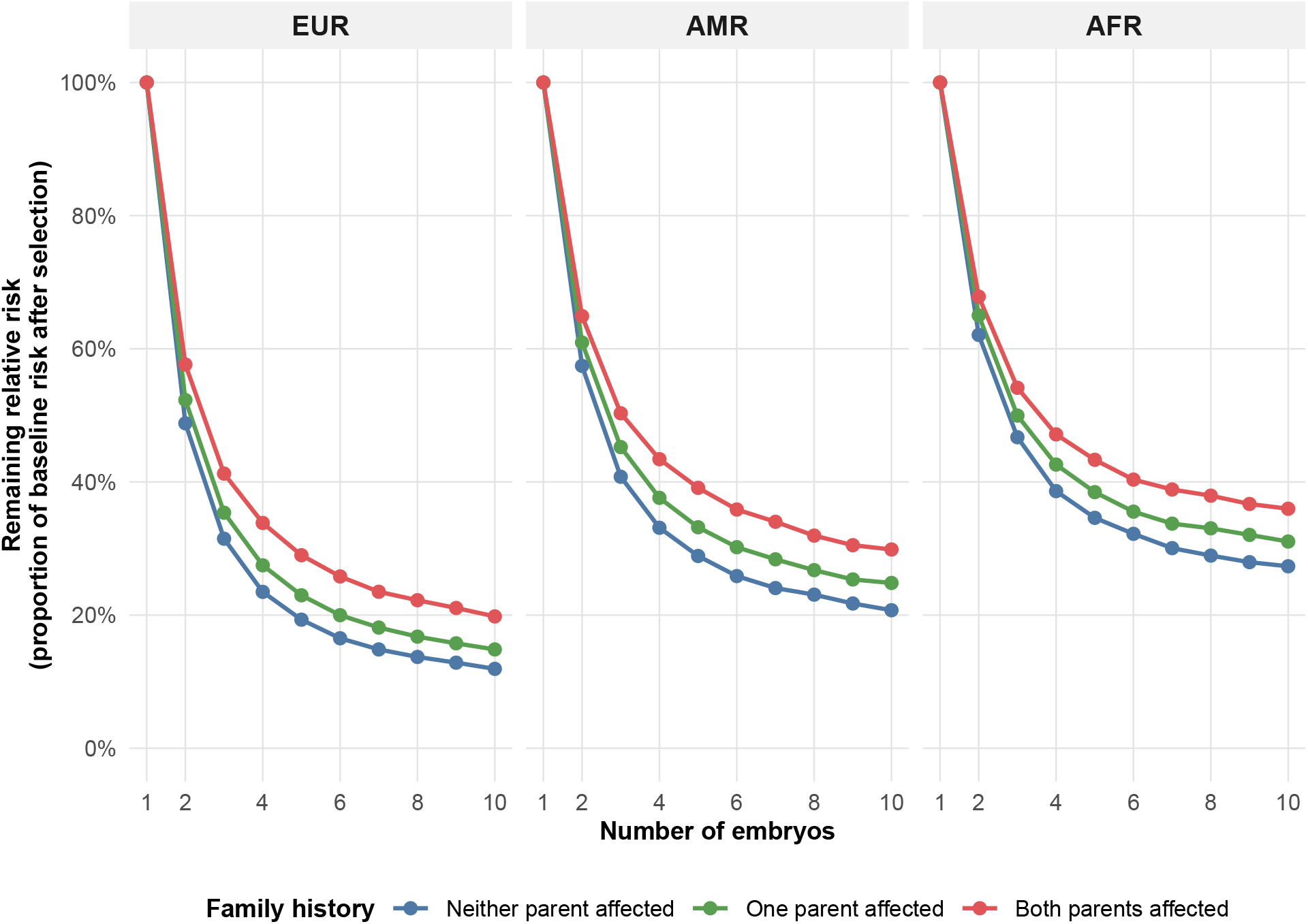
Remaining relative risk after HLA-ARC-based embryo selection across ancestries and family histories. Lines show the proportion of baseline T1D risk remaining after selection of the embryo with lowest HLA-ARC score, as a function of the number of embryos available. Each panel shows a different genetic ancestry. Colors indicate parental disease history: neither parent affected (blue), father affected (green), mother affected (red), and both parents affected (purple).

RRR exhibits diminishing returns as embryo number increases, with marginal gains declining beyond 5–6 embryos. This plateau reflects both sampling properties and biological constraints on the best achievable genotype from parental genetic potential. Still, with 10 embryos available, risk reduction becomes more pronounced, achieving RRR of 85% in EUR, 69% in AFR, and 75% in AMR for the typical case of One parent affected—effectively reducing T1D risk to near-background levels.

The gap between measured-genetics and oracle selection (Tables S1–S3) provides insight into the limits of current genetic knowledge. For families with neither parent affected and 2 embryos available, HLA-ARC achieves 91% of oracle performance in EUR, 79% in AFR, and 85% in AMR. Performance relative to oracle improves for one-parent-affected families (90% in EUR, 77% in AFR, 83% in AMR) and increases further with more embryos available, reaching 97% in EUR, 87% in AFR, and 92% in AMR with 10 embryos. This high efficiency indicates that HLA-ARC captures the vast majority of heritable variance relevant to within-family selection. The ancestry-specific differences in efficiency likely reflect varying contributions of non-HLA polygenic variation and differences in LD structure, with HLA explaining a larger proportion of total genetic variance in EUR populations. Nevertheless, even in AFR populations where efficiency is lowest, HLA-ARC still captures over three-quarters of the theoretically achievable risk reduction.

Our results demonstrate that HLA-ARC enables meaningful risk stratification within families across diverse ancestries. While T1D differs from highly penetrant Mendelian diseases, the disease imposes substantial lifetime burden including daily management, complications risk, and psychological impact, without current prevention strategies. Families with strong T1D history, particularly those already pursuing IVF, may reasonably value this information ^16^. The magnitude of achievable risk reduction varies with family history, ancestry, and embryo number, but remains substantial even in conservative scenarios, with HLA-ARC maintaining relatively consistent performance across populations.

## Discussion

Our study introduces HLA-ARC, a novel T1D genetic risk score that integrates HLA-haplotype calling with functionally informed polygenic scoring using SBayesRC. Previous T1D PGS efforts have taken one of three approaches: not explicitly modeling HLA, relying on a fixed set of known variants genome-wide, or using standard genome-wide PGS approaches such as PRS-CS for genome-wide data. Here we show that bridging these approaches can be advantageous: by modeling the HLA region in a biologically informed manner via direct genotyping of important HLA alleles, and simultaneously applying annotation-aware PGS approaches such as SBayesRC (which includes a much larger set of genome-wide markers for the non-HLA scoring), one can consistently achieve better overall predictive performance compared to published methods. Our analysis highlights the importance of separately modeling HLA and non-HLA components of polygenic risk in T1D, and underscores how different modeling strategies can impact performance across ancestries. When partitioning the HLA-ARC score into HLA and non-HLA components, the choice of standardization critically influences cross-ancestry interpretation of the results. For the HLA component, allele frequency differences across populations represent true sources of genetic variance contributing to disease risk, thus, a global standardization across all ancestries preserves a biologically meaningful scale to have a valid comparison of effect sizes as shown in Table 2. In contrast, non-HLA score variance, which represents the genome-wide component of the score, reflects demographic drift and GWAS ascertainment across populations, warranting within-ancestry standardization to control for these non-causal differences. After pooled scaling of the HLA component, we observe that this portion of the score emerges as the primary and broadly comparable driver of risk across ancestries, capturing shared genetic susceptibility underlying T1D across all ancestries studied.

T1D-MAPS’s flexible framework for direct genotyping of HLA haplotypes, combined with precise LD-mapping and functional annotation-aware genome-wide scoring of non-HLA components of the risk from SBayesRC, provides a more complete assessment of the genetic architecture of T1D, and hence a more robust estimate of the genetic risk across different ancestries. For ex-ample, in the EUR cohort, the HLA-ARC model reached an AUROC as high as *∼*0.91, slightly above the *∼*0.88 achieved using PRS-CS weights, with similar trends in the non-EUR cohorts. Although these performance gains appear modest in magnitude, they were consistently observed, indicating a tangible improvement from PRS-CS to SBayesRC. Additionally, our results align with the broader observation that SBayesRC often outperforms other PGS algorithms by better fine-mapping causal signals ^8^. When predicting rare diseases like T1D, even a small increment in AUROC or variance explained by the PGS can provide meaningful improvements for disease stratification, potentially translating to better identification of high-risk individuals.

While HLA-ARC showed the best performance overall, it is important to note that TA-PS also demonstrated robust cross-ancestry performance and clear improvement over the baseline PRSedm approach. This suggests that non-HLA loci continue to have a meaningful impact on the genetic risk of T1D, and comprehensive inclusion of them into the overall T1D PGS is crucial. However, despite clear improvements in the non-HLA portion of the score, TA-PS still relies on PRSedm to derive the HLA risk component, which means that TA-PS’s HLA scores are not as finely tuned as T1D-MAPS’s direct haplotype-based genotyping and scoring of the HLA region. In contrast to the two methods that use genome-wide data for non-HLA portions of the risk, PRSedm provides the simplest and quickest approach for T1D genetic risk stratification by scoring only 67 risk variants known to have high effect on T1D. Despite the relatively good performance of PRSedm, lack of explicit ancestry tailoring for the HLA region and not using genome-wide data for non-HLA regions, results in an observable performance lag for PRSedm compared to both TA-PS and T1D-MAPS, especially in non-EUR samples. This observation is further reflected in the larger decline in predictive accuracy of T1D PGS scores for AFR and AMR individuals with PRSedm relative to both TA-PS and T1D-MAPS. For instance, PRSedm saw a pronounced drop in AUROC from EUR to AFR individuals, whereas HLA-ARC maintained high discrimination even in non-EUR individuals (0.89 in non-EUR vs 0.91 in EUR), effectively closing most of the cross-ancestry performance gap.

The high predictive accuracy of HLA-ARC across ancestries suggests immediate utility for preimplantation genetic testing for polygenic conditions (PGT-P). Our simulation framework, which models genetic transmission and accounts for unmeasured familial factors, demonstrates substantial risk reduction achievable through embryo selection based on HLA-ARC scores (see Supplementary Methods). Even with just 2 embryos—a worst-case realistic clinical scenario—HLA- ARC-guided selection reduces T1D risk by approximately 51% in EUR, 43% in AMR, and 38% in AFR families where neither parent is affected. Risk reductions are substantial in families with high disease burden: Remarkably, for the typical use case of a couple where one parent suffers from T1D, HLA-ARC-based selection reduces the child’s risk by almost 90% in european populations.

Effective genetic counseling for couples considering PGT-P for T1D requires integration of both specific HLA haplotype information and comprehensive genetic model-based risk estimates for holistic decision-making. While population-level risk predictions inform baseline expectations, individual families benefit from understanding their specific HLA genotypes and how these interact with family history patterns to influence offspring risk. Counselors should emphasize that PGS-based estimates represent probabilistic projections rather than deterministic outcomes, and that even after screening, residual risk remains due to environmental factors and unmeasured genetic variation. Nevertheless, the magnitude of achievable risk reduction represents clinically meaningful impact for couples facing hereditary T1D risk. A holistic counseling approach balances quantitative risk information with family values, reproductive autonomy, and consideration of T1D’s medical manageability relative to other conditions for which PGT is commonly utilized.

We have demonstrated improvements in T1D PGS prediction, but there are several limitations to our study. First, we were unable to perform within-family validation of the proposed HLAARC approach. Such within-family validations are important to confirm that PGS performance is not inflated by population stratification or other confounding factors. However, the AoU dataset was underpowered for this purpose due to its limited sample size and the scarcity of multiplex T1D families. Nevertheless, the genetic architecture of T1D suggests that attenuation of PGS performance from family structure is likely minimal. HLA alleles are the strongest risk factors for T1D, and they exert large effects even within families. For example, siblings who share both high-risk HLA haplotypes have a significantly higher chance of developing autoimmune disease than siblings without them ^17^. This supports the view that our PGS is capturing direct genetic effects rather than potential confounding. Still, as larger cohorts or family-based datasets become available, future work should formally confirm PGS performance within families to further validate this approach.

Second, sample size imbalances across ancestry groups, particularly the limited number of T1D cases of non-EUR ancestry, may have affected the stability of performance estimates. The wide confidence intervals and overlapping performance metrics across methods suggest that some comparisons, such as the gain of HLA-ARC over T1D-MAPS, may not consistently reach statistical significance with current sample sizes. This underscores the need for larger cohorts to definitively distinguish performance differences.

Third, our case-control definitions relied on established EHR-based criteria, which carry a risk of phenotype misclassification, undetected inclusion of atypical diabetes cases, or misclassified controls, all of which could attenuate the apparent predictive power of the PGS. Future work should therefore prioritize external validation in other large biobank cohorts and refine case definitions, ideally leveraging direct clinician-confirmed diagnoses, when available.

Fourth, although our evaluation included EUR, AFR, MA, and AMR cohorts, not enough Asian samples with T1D were available in AoU for a proper analysis. Nonetheless, our framework is broadly generalizable: the major genetic determinants of T1D, including HLA-DR/DQ alleles, and key non-HLA loci, are relevant across ancestral backgrounds, even if the specific risk variants or allele frequencies differ ^1^. For instance, DR3-DQ2 and DR4-DQ8 drive much of the risk in EUR populations, whereas East Asian populations carry distinct high-risk alleles such as HLA-DR9 in Japan ^18^. If our current score were applied directly to East Asian cohorts without re-weighting, performance would likely be attenuated because many EUR-trained variants have different allele frequencies or weaker effect sizes in East Asians, consistent with cross-ancestry portability challenges seen in other complex diseases. However, our proposed framework can recover a substantial amount of accuracy by incorporating population-specific HLA signals through direct genotyping and by recalibrating genome-wide weights with SBayesRC using East Asian reference data. Based on prior PGS transferability studies for complex diseases, we would expect performance close to what we achieve in AMR, and possibly higher than AFR populations ^19^.

Fifth, we did not perform direct HLA allele-calling; instead, we imputed classical DRB1– DQA1–DQB1 alleles using SNP2HLA ^20^. AoU’s current large-scale genomics release is based on short-read WGS, which does not by itself resolve long-range phase across the MHC. While HLA imputation can be quite accurate for common European haplotypes (e.g., *∼*95–97% two-field with the T1DGC panel), it degrades for rarer alleles and across ancestries, introducing non-differential misclassification that will attenuate HLA effect sizes and, by extension, our performance estimates ^20^. In head-to-head studies, short-read WES/WGS allele-calling (e.g., HLA-HD/HLA*LA) is more accurate than imputation and increases discovery power for autoimmune traits, underscoring that even a short-read caller would likely improve our HLA component ^21^. Nevertheless, long-read sequencing (especially PacBio HiFi) enables full-length, fully phased HLA haplotypes with *∼*99.8% per-base accuracy and has been shown to resolve class II ambiguities, which we expect would yield the highest-fidelity DR/DQ haplotypes and sharpen risk stratification in future iterations of our pipeline ^22^. Given the relatively greater importance of the HLA to non-HLA component of HLA-ARC, accurate HLA allele-calling is crucial particularly for maximizing risk prediction accuracy in underrepresented ancestries.

Sixth, our simulation framework models HLA effects additively, which likely provides conservative risk estimates in families carrying multiple T1D-predisposing HLA haplotypes. Substantial evidence indicates synergistic epistasis among risk alleles at the HLA locus, where certain haplotype combinations confer disproportionately elevated risk beyond additive expectations ^**?**^ . For example, the DR3/DR4 heterozygous genotype exhibits an odds ratio substantially exceeding the multiplicative combination of individual DR3 and DR4 effects. This epistatic architecture suggests our additive modeling approach may underestimate the true benefit of selection against high-risk HLA combinations, particularly in families where both parents carry predisposing haplotypes. Future refinements incorporating empirically-derived epistatic effects could further enhance risk prediction accuracy and potentially reveal even greater selection benefits than our current estimates suggest.

The success of our approach in the multi-ancestry AoU cohort demonstrated the feasibility of developing a robust cross-population T1D PGS. However, continued efforts to expand portability of PGS to diverse ancestries and refine modeling strategies will be critical to delivering clinically meaningful risk assessments for T1D worldwide.

## Methods

### Study Cohort

This study used genetic and phenotypic data from the All of Us (AoU) Research Program’s Controlled Tier Dataset Curated Data Repository version 8, available to authorized users on the Researcher Workbench. T1D case and control definitions were based on the validated eMERGE phenotyping algorithm ^23^. The criteria integrate diagnosis codes, medication records, and age of onset to maximize specificity for autoimmune, early-onset T1D while minimizing misclassification from other forms of diabetes. Individuals were classified as T1D cases for the analysis if they satisfied all of the following criteria to ensure high specificity for T1D while minimizing inclusion of secondary atypical diabetes: early-onset T1D while minimizing inclusion of secondary or atypical diabetes:

- At least two recorded T1D diagnosis codes to reduce the risk of miscoding common in EHR data.
- Age at first T1D diagnosis under 40 to enrich for autoimmune T1D and exclude late-onset presentations more consistent with type 2 or atypical diabetes.
- Initiation of outpatient insulin therapy within 365 days of the first T1D diagnosis to capture insulin dependence from disease onset.
- No recorded use of non-insulin diabetes medications prior to insulin initiation, ensuring that treatment history was consistent with T1D rather than type 2 or atypical diabetes.
- No diagnostic codes indicative of cystic fibrosis, cancer, drug-induced diabetes, or other secondary causes of diabetes, to exclude confounding conditions.

Controls were defined using a conservative eMERGE recommended phenotyping algorithm to minimize false negatives and exclude individuals with any potential diabetes-related conditions or medications. Individuals used as controls were required to have:

- At least two least of longitudinal EHR data between the first and last recorded condition, ensuring adequate opportunity to capture any diabetes diagnosis, if present.
- No diagnosis codes for type-1 or type-2 diabetes.
- No prescription for either insulin or non-insulin diabetes medications.
- No diagnosis codes for secondary forms of diabetes or malignant neoplasms.

This strict case-control framework prioritizes specificity over sensitivity, providing a highconfidence T1D phenotype for genetic analyses while minimizing contamination from type 2 or atypical diabetes.

The final cohort included 200 cases and 21,802 controls, with the ancestry distribution detailed in Table 3. For ancestry-specific analyses, only participants of EUR, AFR, and AMR ancestry were retained due to power limitations in other groups.

**Table 3:** Distribution of cases and controls by genetic ancestry in the study cohort. MA includes AFR, AMR, and any other ancestry group that did not reach a threshold required for the analysis.

| Cohort | N_Cases | N_Controls |
| --- | --- | --- |
| EUR | 78 | 7,716 |
| AFR | 54 | 4,029 |
| AMR | 51 | 6,462 |
| MA | 122 | 14,086 |

For the genetic data, we used AoU Allele Count Allele Frequency (ACAF) whole-genome sequencing (WGS) dataset for all subsequent analyses. This dataset includes variant calls from WGS, encompassing both SNPs and INDELs, with a population-specific allele frequency filter of over 1% and an allele count exceeding 100 in any of the computed subpopulations. It comprises approximately 58 million variant sites across *∼*415,000 samples and is optimized for efficient analysis of common genetic variation in AoU by minimizing rare-variant noise and reducing computational burden.

### PGS models

#### PRSedm

The PRSedm approach is an improved version of the GRS2x score, which integrates 67 variants, including those in HLA class I, HLA class II, and non-HLA genome-wide data, partitioned into separate component scores. For each category, a partitioned polygenic score is calculated as the weighted sum of risk alleles using published effect sizes.

More specifically, HLA scores are derived from 36 variants marking HLA haplotypes. Due to imperfect LD between SNPs and true HLA alleles, the method uses imputation and frequency based filtering to assign the most likely haplotype pair, and then converts these into a polygenic distribution. The non-HLA scores are based on the remaining risk variants outside the HLA region. Each allele effect is estimated from the GWAS sources on T1D, with missing variants imputed using reference frequencies under Hardy–Weinberg equilibrium expectations. The combined scores are then obtained by summing the three partitions into a total standardized score. To ensure comparability across studies, PRSedm total score is standardized to a fixed theoretical range using minimum to maximum relative risk normalization. This approach allows separate inspection of HLA and non-HLA contributions, while also providing a single unified score (called GRS2x in PRSedm) for the overall impact of genetic risk on T1D.

#### TA-PS

The TA-PS approach is a simple extension of PRSedm that combines an optimized non-HLA polygenic score by replacing the non-HLA component of PRSedm (which includes only a limited number of markers) with genome-wide weights derived from PRS-CS. The two components of the score are then combined into a unified PGS using a simple linear approach.

#### T1D-MAPS

T1D-MAPS is a recently developed approach that explicitly models HLA and non-HLA components of the T1D risk by directly genotyping HLA alleles and combining them with genome-wide non-HLA scores. For the HLA component, classic HLA alleles are imputed using SNP2HLA approach with a multi-ancestry WGS reference panel. Phased multi-locus haplotypes spanning HLA-DRB1, HLA-DQA1, and HLA-DQB1 are then compared to a reference set of 38 haplotypes derived from a multi ancestry T1D GWAS, with each haplotype assigned a score equal to its log odds of T1D association. In case of ambiguous or unresolved haplotypes, a neutral score of zero is assigned to that haplotype. For the non-HLA component, genome-wide variants summary statistics data from outside the MHC region were used to generate polygenic weights using the PRS-CS method by applying continuous shrinkage prior to approximately 1.2M HapMap3 variants using the EUR 1000 Genomes reference panel.

To construct the overall score, a logistic regression model was fit in the MGB Biobank ^24^ to estimate weights for the HLA and non-HLA components and the resulting coefficients (*β*-HLA and *β*-non-HLA) were then applied to combine the two partitions into a single linear prediction.

#### HLA-ARC

Our proposed approach extends T1D-MAPS by retaining the original partitioned design and explicit modeling of HLA haplotypes, but replacing the PRS-CS based genome-wide component with weights derived from SBayesRC. As in T1D-MAPS, classical alleles at HLA- DRB1, HLA-DQA1, and HLA-DQB1 were imputed using SNP2HLA, then phased into multilocus haplotypes in the cohort. For the non-HLA region, we jointly modeled GWAS summary statistics data with LD from the 1000 Genomes panel and functional annotations (BaselineLD) to shrink SNP effects toward biologically informed priors that account for functional effect of each variant and its LD pattern. Posterior mean effect sizes from SBayesRC were calculated across 7.4 million variants, and individual non-HLA scores were obtained by summing allele dosages weighted by these posterior estimates using the scoring function in plink2. Finally, the combined score was derived by using the same formula used in the original T1D-MAPS publication to yield the integrated predictor. This formulation preserved the strong disease-defining contribution of HLA haplotypes while leveraging the greater modeling capacity of SBayesRC for the genomewide component of the score.

#### Continuous ancestry adjustments for T1D-MAPS and HLA-ARC

To account for continuous genetic ancestry, we used the approach described in T1D-MAPS2, which adjusts an individual’s PGS by weighting the proportion of EUR ancestry using the UK Biobank’s pan-ancestry framework. Individuals with higher EUR ancestry receive greater weight from the PRSedm score, which is optimized for EUR ancestry, while the direct genotyping–based score receives more weight as ancestry diverges from EUR. This formulation allows the score to adapt smoothly across the ancestry spectrum rather than relying on a discrete EUR versus non-EUR classification. By modeling ancestry as a continuous probability instead of a categorical label, the approach can be applied consistently across populations without requiring self-reported or PCA-derived ancestry assignment, thereby reducing bias and improving score portability.

### Evaluation metrics

Model performances were assessed using the area under the receiver operating characteristic curve (AUROC) and the liability-scaled *R*^2^.

AUROC quantifies the discriminative ability of the scores by measuring the probability that a randomly chosen case has a higher score than a randomly chosen control, equivalent to the rank-based separation of cases and controls across thresholds.

The liability-scaled *R*^2^ quantifies the proportion of variance in the unobserved liability to disease explained by the score, given an assumed population prevalence of 0.55% ^25^ for T1D. We developed an HLA haplotype frequency-aware estimator that remains valid under arbitrary predictor distributions (see Supplemental Methods).

Briefly, predicted case probabilities were first obtained in a stratified *K*-fold cross-validation scheme, ensuring out-of-fold predictions for all individuals. Predicted probabilities were recalibrated to the population scale using a prior-odds correction derived from Bayes’ rule:

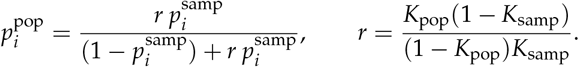

Each population-calibrated probability 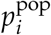 was then mapped to the corresponding latent liability mean under a probit link,

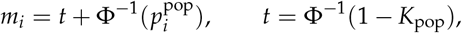

and the variance of *m*_*i*_ provides a direct estimate of the signal variance on the liability scale. The proportion of liability variance explained by the score is therefore

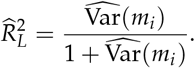

Uncertainty was assessed using 2,000 bootstrap replicates resampling individuals.

### Preimplantation Genetic Testing Simulation

To evaluate HLA-ARC’s utility for preimplantation genetic testing (PGT-P), we developed a simulation framework modeling T1D risk reduction through embryo selection. Full mathematical details are provided in Supplementary Methods.

We modeled individual T1D risk using a liability-threshold framework where disease manifests when total liability exceeds a threshold. Liability comprises four components: (1) measured HLA effects from phased DRB1-DQA1-DQB1 haplotypes with empirically-derived logodds ratios for 38 haplotype combinations, (2) polygenic score effects with ancestry-specific effect sizes (*β*_PGS_: EUR = log(2.3), AMR = log(1.6), AFR = log(1.21)), (3) unmeasured genetic effects 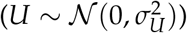, and (4) environmental variation 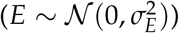.

Variance components were calibrated for each ancestry to reflect empirical heritability estimates (assuming total heritability 85%). For all ancestries, HLA and polygenic score variances were adjusted based on validation performance (see Table 3), with unmeasured genetic variance absorbing the difference to maintain constant total heritability. The baseline parameter *α* was calibrated using Monte Carlo integration over ancestry-specific HLA haplotype frequencies and root-finding with 20-point Gauss-Hermite quadrature assuming a prevalence of 0.55

Ancestry-specific HLA haplotype frequency distributions were derived from 1000 Genomes Project samples. For each superpopulation (EUR, AMR, AFR), we extracted phased DRB1-DQA1- DQB1 haplotype frequencies, mapping each to one of 38 established T1D risk categories or an “other” category with neutral effect. These empirical frequency distributions serve as population priors *p*(*G*) for parental measured genetics when genotypes are unknown—the typical counseling scenario. This approach ensures that simulated risk reductions reflect realistic distributions of parental genetic architectures conditional only on disease history, without requiring actual parental genotyping.

For each simulated family, parental HLA haplotypes were sampled from ancestry-specific frequency distributions and polygenic scores from 𝒩 (0, 1). When parental genotypes are unknown (the standard scenario), we marginalized over latent parental measured genetics by drawing from the population prior *p*(*G*) and reweighting via Bayes’ rule using disease status to obtain *p*(*G* | *D*). This importance sampling approach uses a pre-computed pool of 100,000 candidate genotypes with associated disease probabilities *P*(*D* = 1 | *G*) evaluated via logistic-normal integration, enabling efficient resampling for affected versus unaffected parents.

When parents were affected with T1D, their unmeasured genetic component *U* was drawn from a Bayesian posterior distribution *p*(*U* | *D* = 1, *G*_measured_) computed via discretized grid sampling over 200 points spanning 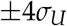 reflecting that affected individuals carry elevated unmeasured genetic risk conditional on their measured genetics.

Embryo genetics followed Mendelian inheritance for HLA (uniform sampling of parental haplotypes) and quantitative genetic transmission for polygenic and unmeasured components. Each embryo’s T1D probability was computed by integrating the liability over environmental variation using 20-point Gauss-Hermite quadrature. To reconcile genetic predictions with empirically observed parent-of-origin effects (2.6-fold higher paternal transmission), we applied stratum-specific adjustment factors: 0.556 for mother-only affected, 1.444 for father-only affected, and 1.0 otherwise.

We simulated 10,000 families for each combination of four parental disease strata (Neither affected, mother affected, father affected, Both affected), three ancestries (EUR, AFR, AMR), and embryo cohort sizes from 2 to 10. For each family, we ran 200 within-family simulations to evaluate variability across possible embryo cohorts. Three selection strategies were compared: (1) random selection (baseline), (2) practical selection based on measured genetics only (HLA + polygenic score, representing clinical implementation), and (3) oracle selection with access to complete genetic information including realized unmeasured components (theoretical upper bound).

## Data Availability

This study used genetic and phenotypic data from the All of Us (AoU) Research Program's Controlled Tier Dataset Curated Data Repository version 8. These data are available to authorized users on the Researcher Workbench. Other data are unavailable.

https://www.researchallofus.org/data/

## Data access statement

This study used genetic and phenotypic data from the All of Us Research Program’s Controlled Tier Dataset Curated Data Repository version 8, available to authorized users on the Researcher Workbench.

## Acknowledgements

We gratefully acknowledge All of Us participants for their contributions, without whom this research would not have been possible. We also thank the National Institutes of Health’s All of Us Research Program for making available the participant data examined in this study.

## Supplemental Tables

**Table S1:** Risk reduction efficiency (AFR).

| Stratum | Embryos | Achieved RRR | Oracle RRR | Efficiency |
| --- | --- | --- | --- | --- |
| Neither affected | 2 | 0.3792 | 0.4775 | 0.7940 |
| Neither affected | 3 | 0.5328 | 0.6371 | 0.8364 |
| Neither affected | 4 | 0.6138 | 0.7158 | 0.8576 |
| Neither affected | 5 | 0.6539 | 0.7569 | 0.8639 |
| Neither affected | 6 | 0.6780 | 0.7829 | 0.8660 |
| Neither affected | 7 | 0.6994 | 0.8040 | 0.8700 |
| Neither affected | 8 | 0.7106 | 0.8168 | 0.8700 |
| Neither affected | 9 | 0.7205 | 0.8281 | 0.8700 |
| Neither affected | 10 | 0.7267 | 0.8364 | 0.8688 |
| One parent affected | 2 | 0.3499 | 0.4533 | 0.7719 |
| One parent affected | 3 | 0.5003 | 0.6127 | 0.8165 |
| One parent affected | 4 | 0.5739 | 0.6879 | 0.8344 |
| One parent affected | 5 | 0.6153 | 0.7315 | 0.8412 |
| One parent affected | 6 | 0.6446 | 0.7614 | 0.8466 |
| One parent affected | 7 | 0.6626 | 0.7812 | 0.8483 |
| One parent affected | 8 | 0.6697 | 0.7928 | 0.8446 |
| One parent affected | 9 | 0.6795 | 0.8046 | 0.8446 |
| One parent affected | 10 | 0.6895 | 0.8152 | 0.8457 |
| Both affected | 2 | 0.3216 | 0.4271 | 0.7529 |
| Both affected | 3 | 0.4585 | 0.5785 | 0.7926 |
| Both affected | 4 | 0.5286 | 0.6539 | 0.8084 |
| Both affected | 5 | 0.5667 | 0.6964 | 0.8138 |
| Both affected | 6 | 0.5965 | 0.7282 | 0.8191 |
| Both affected | 7 | 0.6114 | 0.7482 | 0.8172 |
| Both affected | 8 | 0.6206 | 0.7619 | 0.8145 |
| Both affected | 9 | 0.6331 | 0.7759 | 0.8159 |
| Both affected | 10 | 0.6402 | 0.7854 | 0.8152 |

**Table S2:** Risk reduction efficiency (AMR).

| **Stratum** | **Embryos** | **Achieved RRR** | **Oracle RRR** | **Efficiency** |
| --- | --- | --- | --- | --- |
| Neither affected | 2 | 0.4257 | 0.4989 | 0.8533 |
| Neither affected | 3 | 0.5922 | 0.6653 | 0.8901 |
| Neither affected | 4 | 0.6688 | 0.7392 | 0.9048 |
| Neither affected | 5 | 0.7112 | 0.7798 | 0.9120 |
| Neither affected | 6 | 0.7414 | 0.8082 | 0.9173 |
| Neither affected | 7 | 0.7594 | 0.8257 | 0.9197 |
| Neither affected | 8 | 0.7693 | 0.8367 | 0.9194 |
| Neither affected | 9 | 0.7826 | 0.8485 | 0.9223 |
| Neither affected | 10 | 0.7927 | 0.8578 | 0.9241 |
| One parent affected | 2 | 0.3906 | 0.4683 | 0.8342 |
| One parent affected | 3 | 0.5476 | 0.6294 | 0.8701 |
| One parent affected | 4 | 0.6239 | 0.7048 | 0.8852 |
| One parent affected | 5 | 0.6681 | 0.7481 | 0.8931 |
| One parent affected | 6 | 0.6980 | 0.7769 | 0.8984 |
| One parent affected | 7 | 0.7162 | 0.7953 | 0.9005 |
| One parent affected | 8 | 0.7325 | 0.8109 | 0.9034 |
| One parent affected | 9 | 0.7465 | 0.8240 | 0.9059 |
| One parent affected | 10 | 0.7517 | 0.8307 | 0.9049 |
| Both affected | 2 | 0.3509 | 0.4317 | 0.8128 |
| Both affected | 3 | 0.4969 | 0.5868 | 0.8468 |
| Both affected | 4 | 0.5658 | 0.6588 | 0.8589 |
| Both affected | 5 | 0.6088 | 0.7027 | 0.8665 |
| Both affected | 6 | 0.6414 | 0.7347 | 0.8730 |
| Both affected | 7 | 0.6599 | 0.7546 | 0.8745 |
| Both affected | 8 | 0.6806 | 0.7740 | 0.8794 |
| Both affected | 9 | 0.6950 | 0.7878 | 0.8821 |
| Both affected | 10 | 0.7015 | 0.7959 | 0.8814 |

**Table S3:** Risk reduction efficiency (EUR).

| **Stratum** | **Embryos** | **Achieved RRR** | **Oracle RRR** | **Efficiency** |
| --- | --- | --- | --- | --- |
| Neither affected | 2 | 0.5119 | 0.5624 | 0.9103 |
| Neither affected | 3 | 0.6853 | 0.7311 | 0.9372 |
| Neither affected | 4 | 0.7651 | 0.8053 | 0.9501 |
| Neither affected | 5 | 0.8069 | 0.8436 | 0.9565 |
| Neither affected | 6 | 0.8348 | 0.8687 | 0.9609 |
| Neither affected | 7 | 0.8516 | 0.8837 | 0.9636 |
| Neither affected | 8 | 0.8628 | 0.8942 | 0.9649 |
| Neither affected | 9 | 0.8715 | 0.9021 | 0.9660 |
| Neither affected | 10 | 0.8807 | 0.9101 | 0.9677 |
| One parent affected | 2 | 0.4770 | 0.5302 | 0.8996 |
| One parent affected | 3 | 0.6462 | 0.6972 | 0.9268 |
| One parent affected | 4 | 0.7253 | 0.7722 | 0.9393 |
| One parent affected | 5 | 0.7702 | 0.8137 | 0.9465 |
| One parent affected | 6 | 0.8002 | 0.8411 | 0.9513 |
| One parent affected | 7 | 0.8187 | 0.8581 | 0.9541 |
| One parent affected | 8 | 0.8324 | 0.8706 | 0.9561 |
| One parent affected | 9 | 0.8424 | 0.8799 | 0.9574 |
| One parent affected | 10 | 0.8516 | 0.8880 | 0.9590 |
| Both affected | 2 | 0.4236 | 0.4782 | 0.8859 |
| Both affected | 3 | 0.5875 | 0.6439 | 0.9124 |
| Both affected | 4 | 0.6617 | 0.7169 | 0.9229 |
| Both affected | 5 | 0.7101 | 0.7627 | 0.9310 |
| Both affected | 6 | 0.7420 | 0.7929 | 0.9358 |
| Both affected | 7 | 0.7650 | 0.8145 | 0.9391 |
| Both affected | 8 | 0.7777 | 0.8268 | 0.9405 |
| Both affected | 9 | 0.7893 | 0.8379 | 0.9420 |
| Both affected | 10 | 0.8020 | 0.8491 | 0.9445 |

## Supplement A: Liability-Scale Variance Explained for Non-Normal Genetic Predictors

### Overview

This appendix describes the estimator we use for the proportion of variance explained on the liability scale, 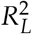, when the binary trait *Y* arises from a normally distributed latent liability *L* via a threshold mechanism, while allowing the genetic predictor(s) to be arbitrarily distributed (including highly non-normal and discrete). This setting is directly relevant to Type 1 Diabetes (T1D), where the HLA region exerts an outsized effect that renders the overall genetic predictor distribution non-Gaussian.

### Liability-threshold model and the target estimand

We assume a general latent variable model

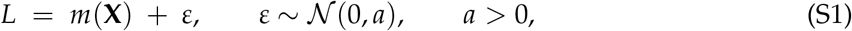

and the binary phenotype

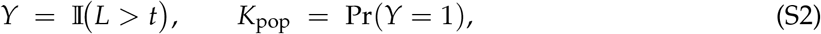

where *m*(**X**) represents the predictor-dependent mean of the latent liability and *a* the residual variance.

The total variance of the latent trait is Var(*L*) = Var { *m*(**X**) } + *a*. The population proportion of liability variance explained by **X** is defined as

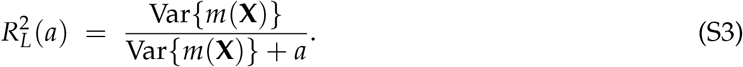

#### Residual-scale standardization

The variance decomposition in (S3) shows that only the *ratio* of Var { *m*(**X**) } to *a* matters. We can therefore fix the residual variance to one without loss of generality:

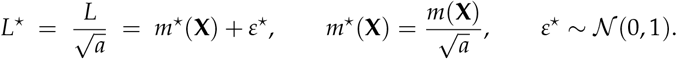

Substituting Var*{m*^*⋆*^ (**X**)*}* = Var*{m*(**X**)*}*/*a* into (S3),

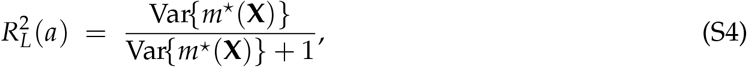

so the choice of *a* cancels algebraically. All expressions henceforth are on this standardized residual scale (*a* = 1).

### Identification of the latent mean from case probabilities

On the standardized residual scale, the model implies

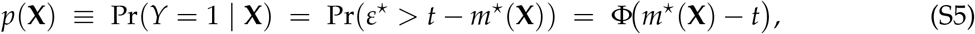

and therefore

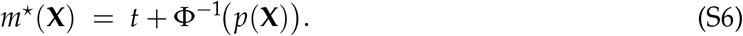

The true threshold satisfying Pr(*Y*=1) = *K*_pop_ satisfies

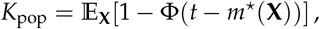

which depends on the entire distribution of *m*^⋆^ (**X**) and is not identifiable from *K* _pop_ a lone. However, the variance in (S4) depends only on the variance of *m*^⋆^ (**X**), not its mean, and is therefore invariant to additive shifts in *t*. For convenience, we adopt the conventional labeling

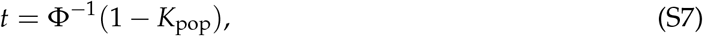

which simply places the population prevalence on the familiar standard-normal scale. This is a choice of origin on the latent axis, not an assumption that *L* itself is standard normal, and it has no effect on the resulting estimate of 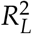.

### Estimator for 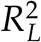

Given fitted or cross-validated case probabilities 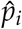, we recover the standardized latent mean per individual using the exact mapping (S6):

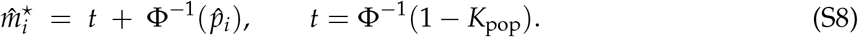

Let 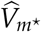 denote the sample variance of 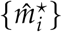. The estimated proportion of liability variance explained is then

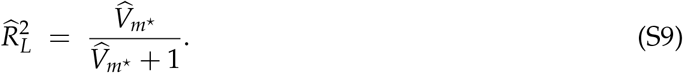

When baseline covariates **C** are included, we compute the analogous variances 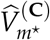 and 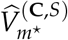 for the baseline and full models, respectively, and report the incremental variance explained:

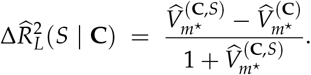

### Correction for case–control ascertainment

In retrospective case–control data, the sampling prevalence *K*_samp_ generally differs from *K*_pop_. Probabilities estimated from such data, 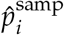, reflect the sample odds rather than population odds. To recover population-calibrated probabilities, we apply a prior-odds correction derived directly from Bayes’ rule.

Let *L*_*i*_ = Pr(**X**_*i*_ | *Y*=1)/ Pr(**X**_*i*_ | *Y*=0) denote the likelihood ratio. Then

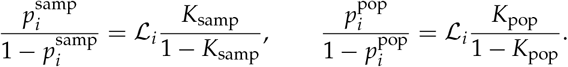

Eliminating ℒ_*i*_ yields

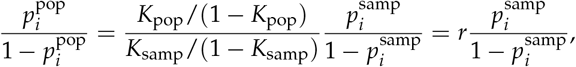

with correction factor

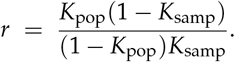

Solving for 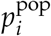 gives

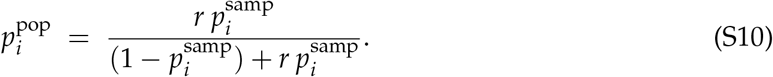

This adjustment shifts only the intercept (prior odds) of the prediction model, preserving its slopes and rank correlations, and ensures the corrected probabilities have marginal mean *K*_pop_.

### Algorithmic implementation

The estimator implemented in `liab_r2_one_score()`proceeds as follows.

#### Algorithm 1

Estimation of liability-scale 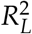 for a single score

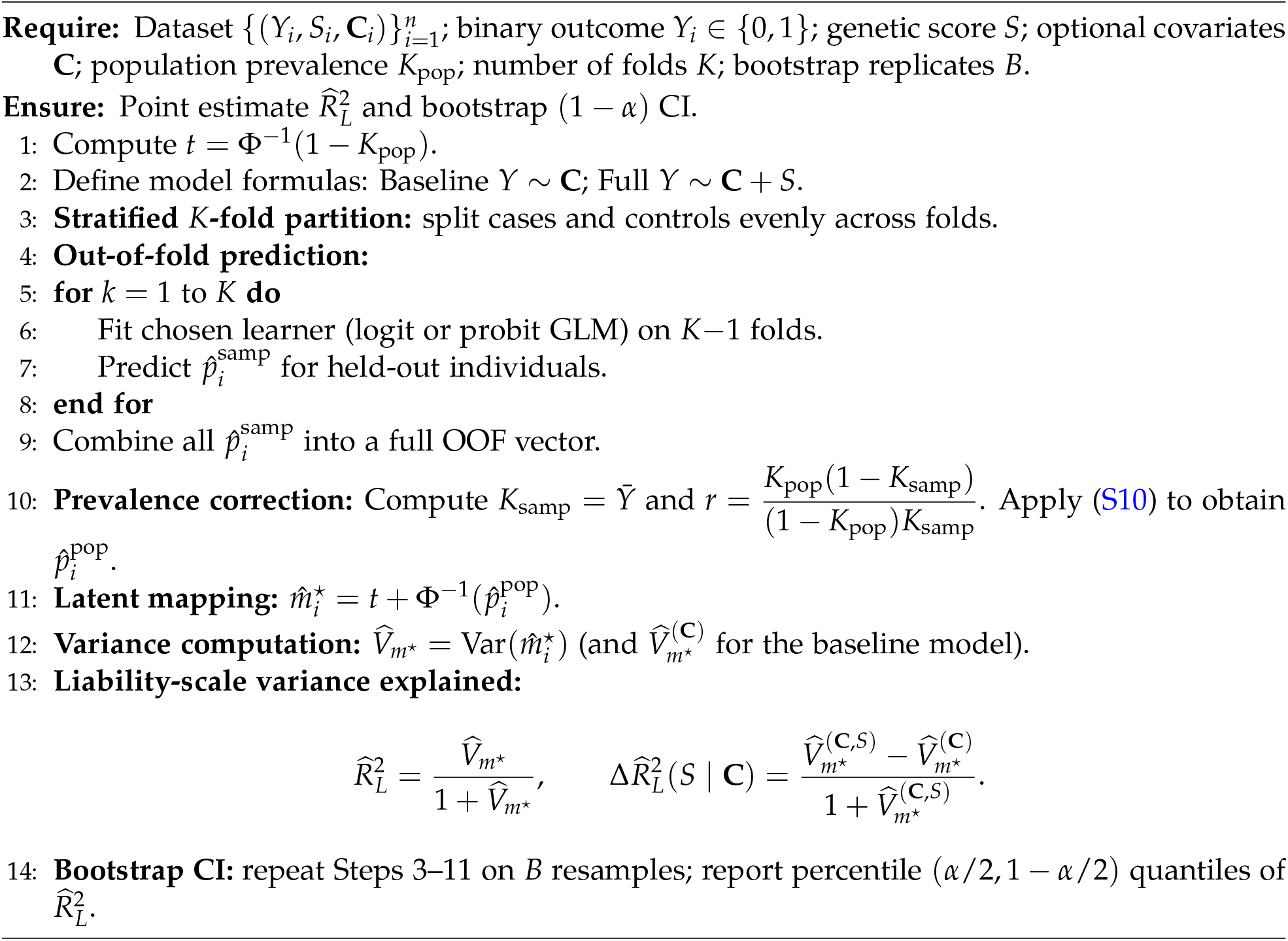

### Uncertainty quantification

We obtain 95% confidence intervals via a nonparametric bootstrap over individuals. Each bootstrap replicate resamples the data with replacement, re-fits the probability models with the same stratified *K*-fold OOF scheme and prior-odds correction, recomputes 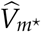 and 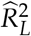, and collects the resulting statistic. The percentile interval between the 2.5th and 97.5th percentiles of the bootstrap distribution is reported.

### Summary

The proposed estimator operates on the standardized residual scale of the liability-threshold model, where only the relative variance of the predictor-dependent mean matters. The threshold *t* = F^*−*1^(1 *− K*_pop_) is adopted purely as a reference labeling of prevalence, not as an assumption about the marginal distribution of *L*. Because 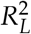 depends solely on the variance of *m*^*⋆*^ (**X**), the scale choice and threshold convention have no effect on the result. This approach yields prevalence-aware, distribution-free estimates of liability-scale variance explained that remain valid even for highly non-normal genetic architectures, such as the HLA-driven component of T1D.

## Supplement B: Simulation of Type 1 Diabetes Risk Reduction Through Embryo Selection

### Overview

This document presents a simulation framework for estimating Type 1 Diabetes (T1D) risk reduction achievable through genetic screening and embryo selection in IVF. The implementation employs a two-stage approach that separates genetic transmission from non-genetic affected parent effects (which remain constant within families).

### Mathematical Model

#### Liability-Threshold Model

T1D risk is modeled using a liability-threshold framework where disease manifests when total liability *λ* exceeds a threshold. For an individual with genetic and environmental components:

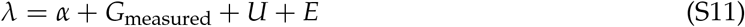

where:

- *α*: baseline log-odds, calibrated to match population prevalence
- *G*_measured_ = *β*_HLA_(*H*) + *β*_PGS_ · *S*: measured genetic effects
- 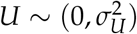: unmeasured genetic liability
- 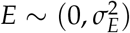: environmental noise

The measured genetic component comprises:

- **HLA effects**: 38 haplotype combinations with empirically-derived log-odds ratios
- **Polygenic score**: Non-HLA genetic risk with *β*_PGS_ = log(2.3) per standard deviation (EUR, modify according to validation results for other ancestries).

#### Risk Calculation via Integration

For an embryo with realized genetics (both measured and unmeasured), the probability of T1D integrates over environmental variation:

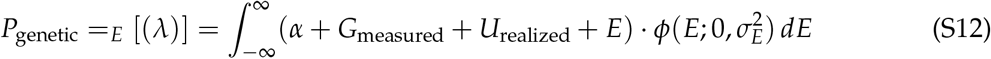

This integral is computed using 20-point Gauss-Hermite quadrature:

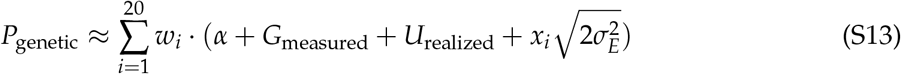

where (*x*_*i*_, *w*_*i*_) are the Gauss-Hermite nodes and weights.

#### Variance Decomposition

The liability-scale variance components reflect empirical heritability estimates as given in the main text.

#### Genetic Transmission Model

For parents with genetic profiles (*G*_*M*_, *U*_*M*_) and (*G*_*F*_, *U*_*F*_), embryo genetics follow:

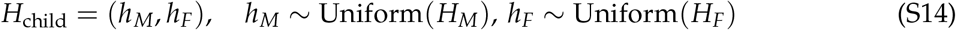

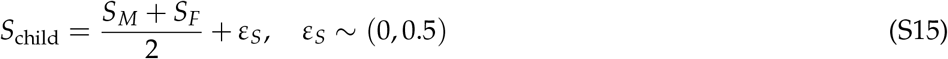

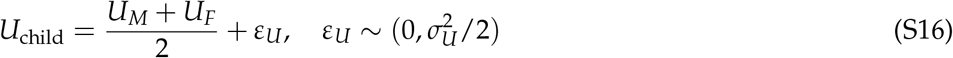

The segregation variances follow from quantitative genetic theory under random mating. Importantly, each embryo receives a unique realization of *ε*_*U*_, creating variation in unmeasured genetics across siblings.

#### Posterior Distribution for Affected Parents

Following Bayes’ theorem, the unmeasured genetic component for an affected parent follows:

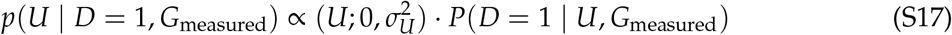

where the likelihood integrates over environmental variation:

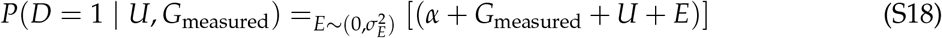

The implementation samples this posterior using a discretized grid with 200 points over *±*4*σ*_*U*_.

### Risk Reduction given known Parental Genotypes

#### Empirical Transmission Patterns

Observed T1D transmission risks show striking parent-of-origin asymmetry. Offspring of affected mothers have a 2.5% risk of developing T1D, compared to 6.5% for offspring of affected fathers, regardless of offspring sex. Research suggests this 2.6-fold higher paternal transmission risk cannot be explained by genetic factors alone. To reconcile genetic predictions with empirical risks, we apply stratum-specific adjustments:

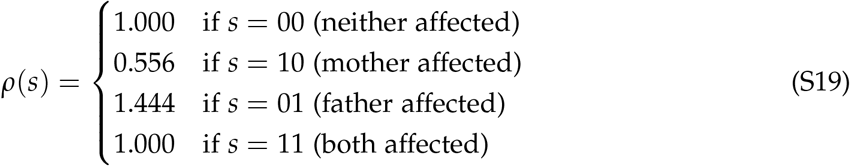

These factors:

- Preserve the average risk in discordant cases: (0.556 + 1.444)/2 = 1.0
- Match the empirical ratio: 1.444/0.556 = 2.6
- Apply uniformly within families (do not affect selection)

#### Final Risk Calculation

The complete risk for a child from family stratum *s* with genetics *G* is:

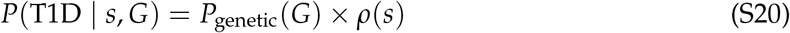

This two-stage approach ensures genetic variation drives selection while absolute risks match empirical data.

### Simulation Algorithm

#### Alpha Calibration

The baseline parameter *α* is calibrated to achieve population prevalence of 0.55%:

1: Initialize *α*_0_ = (0.0055)

2: Define objective: *f* (*α*) = [*P*_population_(*α*)]*−* 0.0055

3: Solve *f* (*α*) = 0 using root-finding with:

- Monte Carlo sampling over HLA frequencies
- Integration over PGS distribution
- Gauss-Hermite quadrature for *U* and *E*

#### Embryo Selection Simulation

For a family with characteristics (*D*_*M*_, *D*_*F*_, *H*_*M*_, *H*_*F*_, *S*_*M*_, *S*_*F*_):

#### Key Distinction: Practical vs Oracle Selection

The critical difference between selection strategies:

- **Practical selection**: Observes only *G*_measured,*j*_ (HLA + PGS), cannot see the realized unmeasured genetics *U*_*j*_ that varies between embryos
- **Oracle selection**: Observes the complete risk *P*_*j*_ which incorporates the specific realized value of *U*_*j*_ for each embryo
- **Selection efficiency**: The ratio of practical to oracle performance reflects the information loss from unobserved genetic variation

### Risk Reduction Given Unknown Parental Genotypes

#### Unknown Parental Genotypes as Latent Variables

Let *D∈ {*0, 1} be a parent’s affection status (T1D). When *G* is unobserved, the correct parental prior for measured genetics is the population distribution *p*(*G*) induced by the ancestry-specific HLA haplotype frequencies and *S∼ N*(0, 1); conditioning on *D* updates this via Bayes:

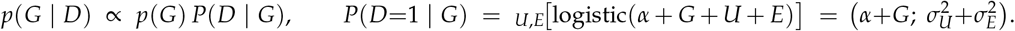

We evaluate *P*(*D*=1 | *G*) with the same Gauss–Hermite integral used elsewhere. The posterior for the unmeasured component *U* given *both D* and *G* remains the grid-based update described in the main text (affected/unaffected parents sample from *p*(*U* | *D, G*)).

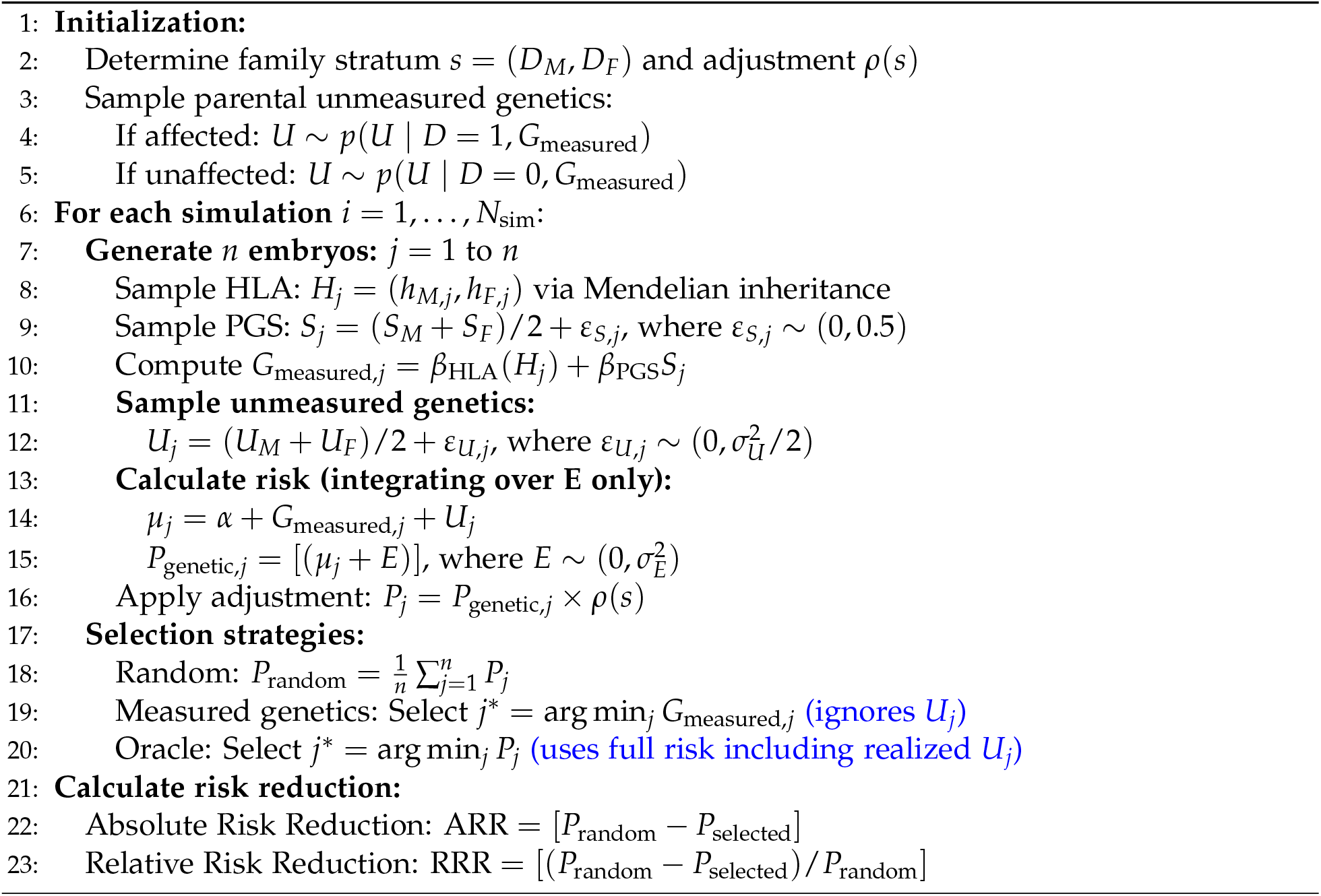

#### Expected Selection Benefit by Family History

For a given stratum *s* = (*D*_*M*_, *D*_*F*_) and *n* embryos, define the within-family (oracle) risk of embryo *j* as *P*_*j*_ = *P*(T1D | *s, G*_*j*_, *U*_*j*_) and the random-selection baseline 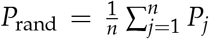. The practical selector observes only *G*_*j*_ and chooses 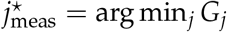; the oracle chooses 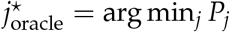.

The *expected* absolute and relative risk reductions *by history alone* average over the unknown measured genetics of both parents:

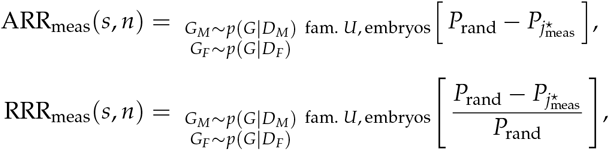

with analogous definitions for the oracle. The inner expectation is exactly the family-level simulation in the main algorithm; the outer expectation marginalizes the unobserved parental measured genetics via *p*(*G* | *D*).

#### Computation

##### Population prior for *G*

We obtain *p*(*G*) by sampling parental haplotypes from the ancestry-specific HLA frequency vector and drawing *S ∼ N*(0, 1); then *G* = *β*_HLA_(*H*_1_, *H*_2_) + *β*_PGS_*S*. Given a large candidate pool 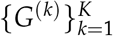 from *p*(*G*), we approximate *p*(*G* | *D*) by self-normalized importance resampling with weights *w*^(*k*)^ ∝ *P*(*D* = 1 | *G*^(*k*)^) (or 1*−P*(*·*) for *D* = 0), where *P*(*D* = 1 | *G*) is evaluated by the logistic–normal integral with variance 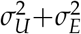.

##### Inside-family simulation

Conditional on the resampled (*G*_*M*_, *G*_*F*_), we reuse the existing pipeline: (i)sample *U*_*M*_, *U*_*F*_ from *p*(*U D, G*) on a fixed grid, (ii) generate *n* embryos by Mendelian HLA plus segregation for PGS and *U*, (iii) compute per-embryo risks by integrating over *E* only, (iv) apply *ρ*(*s*), and (v) evaluate random/practical/oracle selection.

##### Alpha and numerics

*α* is calibrated once per ancestry to match the target prevalence *K* using the same root-finding objective as before: Monte Carlo over HLA/PGS and Gauss–Hermite for *U*+*E*. The same Gauss–Hermite machinery is reused for the *P*(*D* = 1 | *G*) integral.

## Notes

### Competing Interest Statement

Herasight, Inc employs M.A., S.M., I.D., J.A., R.M., J.H.L., D.S., and T.W. M.C. is CEO of Herasight, Inc. All authors hold equity in Herasight, Inc.

### Funding Statement

Herasight, Inc funded this study.

### Author Declarations

The IRB of All of Us gave ethical approval for this work.

### Summary of Updates

Herasight, LLC was changed to Herasight, Inc in the competing interests statement.

